# Daily movement behaviours and cognition in mid-life: A cross-sectional compositional analysis of the 1970 British Cohort Study

**DOI:** 10.1101/2022.07.06.22277309

**Authors:** JJ Mitchell, JM Blodgett, S Chastin, BJ Jefferis, G Wannamethee, M Hamer

**Affiliations:** Department of Primary Care and Population Health, London, UK; Upper Third Floor UCL Medical School (Royal Free Campus), Rowland Hill Street, London, UK. NW3 2PF; Institute for Applied Health Research, School of Health and Social Care, Glasgow Caledonian University, Glasgow G4 0BA, UK; Institute of Sport Exercise & Health, Division of Surgery and Interventional Sciences, Faculty Medical Sciences, University College London, London, UK

**Keywords:** Accelerometer, isotemporal substitution, moderate and vigorous physical activity, movement, compositional analysis, sedentary behaviour, sleep

## Abstract

**IMPORTANCE:** Movement behaviours (e.g. sedentary behaviour (SB), moderate and vigorous physical activity (MVPA), light intensity physical activity (LIPA) and sleep) are linked to cognition, yet the relative importance of each component is unclear, and not yet explored with compositional methodologies.

**OBJECTIVE:** To examine how time spent in one behaviour (e.g. SB, MVPA, LIPA, sleep) relative to all others is associated with overall cognition, including executive function and memory.

**DESIGN, SETTING, AND PARTICIPANTS:** The 1970 British Cohort Study (BCS70) is an ongoing prospective birth cohort study of adults born in England, Scotland and Wales in a single week. At age 46, participants wore an accelerometer device and completed cognitive screening. Linear regression was used to examine cross-sectional associations between movement behaviours and cognitive scores, using a compositional approach. Isotemporal substitution was performed to model the effect of reallocating time between components of daily movement on cognition.

**EXPOSURES:** A thigh-mounted activPAL accelerometer device was worn without removal for one week. Daily time in SB, MVPA, LIPA and sleep were derived using a validated processing algorithm.

**MAIN OUTCOMES AND MEASURES:** Standardised scores of executive function (letter cancellation and verbal fluency), memory (immediate and delayed wordlist recall) and a composite score were derived from computer administered cognitive tasks.

**RESULTS:** The sample comprised 4,481 participants (52% female). Time in MVPA relative to SB, LIPA and sleep was positively associated with cognition after adjustments for education and occupational physical activity, but additional adjustment for health status attenuated associations. SB relative to all other movements was robustly positively associated with cognition. Modelling time reallocation between components revealed a higher cognition percentile after MVPA theoretically replaced just 9 minutes of SB (+1.31; 95% CI: 0.09, 2.50), 7 minutes of LIPA (+1.27; 0.07, 2.46) or 7 minutes of sleep (+1.20; 0.01, 2.39).

**CONCLUSIONS AND RELEVANCE:** Relative to time spent in other behaviours, more time spent in MVPA and SB was associated with higher cognitive scores. Displacement of MVPA time, given its smaller relative amount, appear most deleterious. Efforts should be made to preserve MVPA time, or reinforce it with time taken from other behaviours.

## Introduction

Physical inactivity remains a principal cause of early morbidity and mortality globally^1-3^ and is implicated in numerous disease pathways^4,5^. Despite the known health effects of physical activity (PA), only recently have studies explored the positive impacts of PA on cognition^6-18^. PA engagement has been linked to the building of cognitive reserve which might help delay the onset of cognitive decline in later life^9,10,14,19-26^. However, all aspects of PA including intensity and volume decrease throughout the life course^18^ with significant drops at critical periods, which may therefore have consequences for cognition later in life^27-32^. As such, disentangling the most important components of PA for cognition remains a pressing question. Furthermore, evidence regarding the magnitude of this association remains under scrutiny^33^, especially outside of later life, and in the years immediately preceding cognitive decline^33,34^.

Evidence from adults in midlife is scarce. Those few studies using objective-measures typically examine only one movement intensity whilst adjusting for another ‘*opposing’* movement intensity and report varied results as to the most important facets of PA^9,35^. One systematic review of such studies report greater total PA and greater moderate and vigorous PA (MVPA) as being most beneficial for cognition^35^. Only two studies have explored the associations of movement behaviours and cognition using more robust, time-exchange methodologies; one in older adults^36^ and one in mid-life^37^. Both cite MVPA as most favourable for global cognition. Both studies however do not capture sleep time which is the final and typically largest component of the day. Modelling the contribution of sleep is acutely relevant when examining cognition, as it is known to be disrupted in adults with cognitive impairment^38-42^ and is a major confounder of test performance.

Compositional data analysis approaches are well established statistical methods for examining multivariate, finite quantities^43-46^. This methodology is increasingly utilised to model daily movement as traditional approaches fail to account for the co-dependency of each component of 24-hour movement, including sleep time. Utilising compositional approaches to explore the relationship of PA and cognition addresses a gap in current literature by examining PA in the context of *all* components of the day in a ‘closed system’^47^.

Given the emergence of compositional methodologies for analysing movement data, and the scarcity of evidence of the critical components of daily PA for cognition in midlife, this study aimed to *(i)* assess the associations of different components of daily movement and participant’s overall cognition (including memory and executive function) using a compositional approach, (*ii)* understand the relative importance of each daily movement intensity for cognition by examining the effect of time reallocation between behaviours^48,49^.

## Methods

### Participants

The 1970 British Cohort Study (BCS70) consists of individuals born within one week of another in England, Scotland and Wales in 1970 and followed up regularly throughout childhood and adulthood^50^. This analysis utilises data from the follow-up at age 46 collected in 2016–2018 (*N=8,581*). The age-46 sweep involved biometric measurements, completion of health, demographic, and lifestyle questionnaires, and invitation to wear a thigh-mounted accelerometer device for one week. Participants provided informed consent prior to participation and ethical approval was obtained from NRES Committee South East Coast - Brighton & Sussex (Ref 15/LO/1446).

Inclusion criteria were restricted to participants who consented to wear an accelerometer and returned the device with sufficient wear time (minimum of 1 day with 10 hours uninterrupted wear time) as well as provided data on all measures relevant to this study (eFigure 1 in the Supplement)^51^.

### Measures

#### Objective measures of PA, sedentary Behaviour and Sleep

PA, SB and sleep time were measured using a thigh-mounted accelerometer device worn continuously for up to seven days including for sleeping and bathing activities (activPAL3 micro; PAL Technologies Ltd., Glasgow, UK). The wear protocol has previously been described and validated against self-reported PA from participant wear-diaries^52-55^. The accelerometer device uses a thigh inclination technique that is more sensitive to detecting different forms of SB than wrist and hip-worn devices^55,56^. MVPA time was derived using a step cadence of ≥100^57^. Light intensity PA (LIPA) was derived as the residual from the total activity, subtracting MVPA. SB was defined as non-sleep time spent sitting or lying, based on posture. Lastly, sleep time, or ‘time in bed’ was considered as the longest reclining bout between noon and noon each day (min ≥2◻hours) or any long bouts lasting ≥5◻hours, and has been shown to be accurately distinguished from bouts of prolonged non-wear^58^.

#### Primary Outcome

Participants undertook computer-administered tests of memory (immediate and delayed word recall tasks) and executive function (verbal fluency and two-letter cancellation tasks). These tools have been widely and routinely used in epidemiological studies to measure cognition^33,59-63^. Immediate and delayed memory tasks involved asking for recall of a list of 10 words over a 2 minute period before the retrieval of these words again after a substantial delay. The word list was randomized between participants from a set of four lists. Immediate and delayed recall was scored as the total number of words recalled correctly in any order. Executive function tasks included a verbal fluency task which scores the number of animals participants could name in 1 minute, while processing speed and accuracy tests involved a 2-letter cancellation task. This task involves participants screening a grid of letters line-by-line, and crossing out as many “P’s” and “W’s” as they identify in 1 minute. Processing speed scores the total number of letters screened while processing accuracy scored the number of letters missed. Z-scores were derived for each test for the analytic sample. Z-scores were summed for each domain (memory and executive function individually) and also summed to produce an overall composite score for cognition^59,64^.

#### Covariates

Covariates were selected a priori based on previous literature and encompassed sociodemographic factors, health factors and lastly risky health behaviours^33-35^.

### Sociodemographic Covariates

Age was included as a continuous measure as the difference in months between participant date of birth on study entry and date of completion of the survey. Sex, also derived from the study entry survey, was coded as female vs male. Lastly, marital status was included as a trichotomous variable with groups: ‘*unmarried’*, ‘*separated, divorced or widowed’* and *‘married or a civil partner’*.

### Highest Educational Attainment

Education was categorised by highest academic attainment by the age 46 follow-up. Participants were categorised as none: *No formal academic qualifications*, lower: *GCSE or O-level (typically attained at aged 16)*, middle: *A-level or regional equivalent (typically attained at aged 18)*, upper: *Diploma or degree level*, and highest: *higher degree*.

### Occupational Physical Activity

Occupational PA was measured using a single item question derived from the EPIC-Norfolk PA questionnaire^65^. Participants were asked, ‘*What is the best corresponding type and amount of physical activity involved in your work?’* with responses: ‘*Sitting occupation’, ‘Standing occupation’, ‘Physical work’, ‘Heavy manual work’, and* ‘*Not working*’.

### Disability

Disability was measured using the EU Statistics on Income and Living Conditions abbreviated measure of disability^66^. This was categorised as ‘*No long-standing health condition*’, ‘*to some extent*’, or ‘*severely hampered’*.

### Body Mass Index (BMI)

Body weight was measured using the Tanita electronic scale, and height was measured using standard protocols. BMI (calculated as; Body mass (*kg*)/height (*m*)^2^) was included as a continuous variable. Where nurse measured BMI was missing (n=66) this was imputed based on self-reported weight and height.

### Psychological Distress

Psychological distress was measured using the Malaise inventory, a validated 9-question scale with a cut-point of 4 indicating psychological distress^67^. Participant answers were dichotomised as above/equal to this cut-point vs below.

### Health Behaviours

Alcohol consumption was categorised as ‘abstinent’, ‘irregular or regular non-risky’, and ‘risky’ (consuming ≥14 weekly units) according to the Alcohol Use Disorders Identification Test scale^68^.

Smoking status was categorised as, *‘never’, ‘ex-smoker’*, ‘*less than daily’* and ‘*daily’*.

#### Statistical Analysis

T-tests and chi-squared tests were used to compare the characteristics of participants who fulfilled eligibility criteria (analytic sample) with those in the excluded sample. Initial analyses explore the differences of participant’s four-part compositions (average proportions of the day spent in MVPA, LIPA, SB and sleep) by quartile, using MANOVA Pillai’s trace test. An isometric log-ratio transformation (ILR) approach was then used to model the association of cognition and daily movement compositions^45,46,69,70^. The isometric log-ratio transformation for compositions of *n* components produces *n*-1 ILR-coordinates which, together, account for all daily movement. Unadjusted linear regression models estimated the associations between ILR coordinates of participant’s PA habits and composite cognition z-scores. The first coordinate in each model was reported as this reflects the association of one component relative to all others with the cognition outcome. Adjustments for potential confounders and mediators were made in two steps: (i) sociodemographic factors including age, sex, education and occupational PA; (ii) health and lifestyle factors including BMI, disability, psychological distress, and risky health behaviours. Next, using the unadjusted model, minute-by-minute isotemporal substitution was performed to model the resultant change in cognition score when time was reallocated from one component of daily movement into another (e.g. MVPA into SB) around the sample mean, whilst holding the other two constant. The point at which Z-scores showed substantial positive or negative change was defined as the point at which the modelled z-score’s 95% CI no longer overlapped with the mean. This was then converted into the corresponding percentile and reported as the improvement in percentile. All analyses were conducted using R studio (RStudio team, Boston, MA, 2020).

#### Sensitivity Analysis

All analyses were repeated with participant’s composite memory z-scores and composite executive function z-scores to explore the differential relationship between PA and these aspects of cognition.

Lastly, we tested a priori hypotheses that the relationship between PA and cognition may differ depending on educational attainment, sex, and occupational PA. As such, we investigated interactions between each of the covariates and the ILR coordinates in the initial unadjusted model. Stratified analyses were then conducted in the case of a significant interaction effect at the level of α<0.01.

## Results

### Participant Characteristics

The analytic sample comprised 4,481 participants (eFigure 1 in the Supplement) aged 46 years, principally female (52% Female, N=2347), mostly married (66%, N=2,954) and high educational attainment (43% attained A-levels or above (typically attained at 18 years or older), N=1,919; Table 1). The majority of participants’ alcohol consumption was occasional or non-risky (68%, N=3,033) and never smoked (50%, N=2,260). Those 2,959 participants who did consent to accelerometer-wear but were excluded due to device error, insufficient wear time or failing to provide relevant covariates were proportionally more male *(p*<0.001) and higher BMI (*p*<0.001) but otherwise did not differ significantly from the analytic sample (Table 1). Participants spent an average of 46 minutes (*m*) in MVPA, 5 hours (*H*) 35m LIPA, 9*H* 20*m* of SB and 8*H* 17*m* sleeping (eFigure 2 in the Supplement). All raw cognitive tests were normally distributed and showed no evidence of floor or ceiling effects, except for processing accuracy, in which participants routinely missed none or few letters (eFigures 3,4 in the supplement). Median processing speed was 356 (286-403) letters processed and 3 missed (Q1,Q3: 1, 6). Mean verbal fluency was 24.0 animals named (SD=6.1). Mean immediate recall was 6.7 words (SD=1.4) and delayed recall was 5.6 words (SD=1.8) (table 2).

**Table 1.** Sample demographics and bias analysis.

| Sociodemographic Characteristics |  | Included Sample | Excluded Sample | Comparison |
| --- | --- | --- | --- | --- |
|  |  | N=4,481 | N=2,959 | p-value |
| Age | Mean, SD | 47 (0.6) | 47 (0.7) | 0.072 |
| Sex (Female) | % (N) | 52% (2347) | 50% (1483) | 0.060 |
| Married | % (N) |  |  |  |
| Unmarried |  | 19% (836) | 20% (553) | <0.001* |
| Divorced, separated or widowed |  | 15% (691) | 18% (520) |  |
| Married or in civil partnership |  | 66% (2954) | 62% (1747) |  |
| Highest Educational Attainment | % (N) |  |  | <0.001* |
| No formal qualifications |  | 25% (1132) | 32% (917) |  |
| GCSE or equivalent |  | 32% (1430) | 30% (869) |  |
| A-level or equivalent |  | 6% (253) | 6% (164) |  |
| Undergraduate degree or equivalent |  | 31% (1407) | 27% (765) |  |
| Higher degree |  | 6% (259) | 5% (143) |  |
| Occupation Type | % (N) |  |  | <0.001* |
| No occupation |  | 8% (377) | 12% (291) |  |
| Sitting |  | 51% (2274) | 47% (1108) |  |
| Standing |  | 14% (629) | 14% (324) |  |
| Physical work |  | 22% (1001) | 23% (547) |  |
| Heavy manual work |  | 5% (200) | 4% (94) |  |
| Health Status & Behaviours |  |  |  |  |
| Disability status (EU-SILC) | % (N) |  |  | <0.001* |
| No EU-SILC long-standing health condition |  | 85% (3811) | 80% (2379) |  |
| EU-SILC classification, disability to some extent |  | 10% (456) | 12% (342) |  |
| EU-SILC classification severely hampered |  | 5% (214) | 7% (236) |  |
| Psychological distress (Abbrev. Malaise Scale) | % (N) |  |  | <0.001* |
| High malaise (4+) |  | 17% (768) | 21% (520) |  |
| Alcohol Consumption (weekly units) | % (N) |  |  | <0.001* |
| None |  | 10% (438) | 12% (360) |  |
| Occasional (<14) |  | 68% (3033) | 63% (1827) |  |
| Regular risky (>14) |  | 22% (1010) | 25% (726) |  |
| Smoker Status | % (N) |  |  | <0.001* |
| Never |  | **50% (2260)** | **44% (1310)** |  |
| Ex-smoker |  | **32% (1146)** | **32% (933)** |  |
| Less than daily |  | **5% (215)** | **6% (175)** |  |
| Daily |  | **12% (560)** | **18% (541)** |  |
| **BMI** | **Mean, SD** | **28.4 (5.6)** | **29.0 (6.2)** | **<0.001\*** |

\*Significant differences between included and excluded sample characteristics presented at the level $\alpha \leq 0.05$ as indicated by t-tests and Pearson's Chi-Squared ( $\chi^2$ ) test.

**Table 2.** Raw participant cognitive test scores and comparison with excluded sample.

|  | **Analytic Sample**<br>**N = 4,881**<br>**Median (IQR)** | **Excluded Sample**<br>**N= 2,959**<br>**Median (IQR)** | **p-value** |
| --- | --- | --- | --- |
| Processing Speed | 335 (117) | 331 (119) | 0.550 |
| Processing Accuracy | 3 (5) | 3 (4) | <0.001* |
| Verbal Fluency | 24 (8) | 23 (8) | <0.001* |
| Recall (immediate) | 7 (2) | 7 (1) | <0.001* |
| Recall (delayed) | 6 (3) | 5 (3) | <0.001* |

\*Significant differences between included and excluded sample characteristics presented at the level $\alpha \leq 0.05$ as indicated by t-tests and Pearson's Chi-Squared ( $\chi^2$ ) test. Processing accuracy is the count of errors (missed letters) on 2-letter cancellation task and is inversely scored.

Participant time in each movement intensity differed significantly between quartiles of composite cognition scores (*p*<0.001, MANOVA Pillai’s trace test, figure 1). Compared to the sample mean, participants in the upper two quartiles of cognition spent greater time in MVPA and SB and had less sleep on average than those in the lower two cognition quartiles, yet the lowest quartile of cognition had the highest proportion of LIPA.

**Figure 1.**
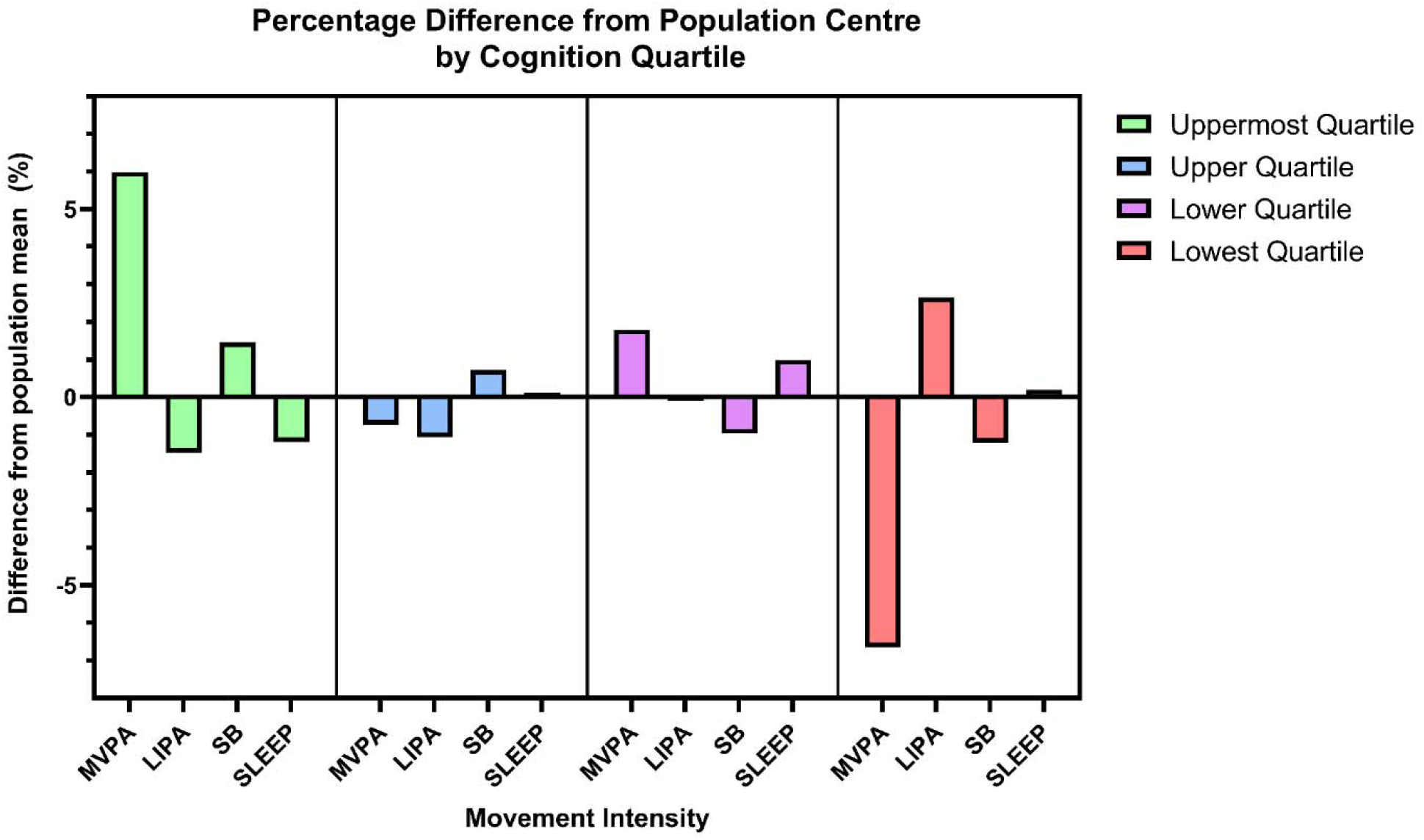
Participant movement profiles by cognition quartile. Percentage difference of compositions of daily movement from the sample compositional mean, stratified by composite cognition quartile (quartile 1; highest cognition – quartile 4; lowest cognition). Significant differences in quartile movement compositions were identified (MANOVA Pillai’s trace test, *p* < 0.001).

### Associations of Daily Compositions and Cognition

Linear regressions revealed a positive association between MVPA relative to all other behaviours, and cognition z-scores (eTable 1 in the supplement). SB, relative to all other behaviours was also positively associated with cognition. These associations persisted with adjustment for sociodemographic factors including age, sex, education, marital status, and occupational PA. The relationship between cognition and MVPA relative to other behaviours was fully attenuated after further adjustment for disability, smoker status, risky alcohol consumption, BMI and psychological distress. However, SB relative to all other movement remained positively associated with cognition after full adjustment. Conversely, more time spent in LIPA or sleep relative to all behaviours was inversely associated with cognition. However, only more time spent in sleep relative to other behaviours remained significantly, inversely associated with cognition in the fully adjusted model.

### Isotemporal Substitution Analysis

To better understand the joint associations of these behaviours with cognition we modelled the change in cognition z-scores (converted to change in percentile) associated with different compositions of the day relative to the sample’s mean composition (*46m* MVPA, *5H 35m* LIPA, *9H 20m* SB, *8H 17m* sleep). Using the unadjusted model, we performed these reallocations minute-by-minute from one of component into another whilst holding the other two constant. Increased cognition percentile was seen after 9 minutes of SB was replaced with (9 min) MVPA (+1.31; 95% CI: 0.09, 2.50; figure 2), 7 minutes of LIPA was replaced with MVPA (+1.27; 0.07, 2.46) or 7 minutes of sleep was replaced with MVPA (+1.20; 0.01, 2.39).

**Figure 2.**
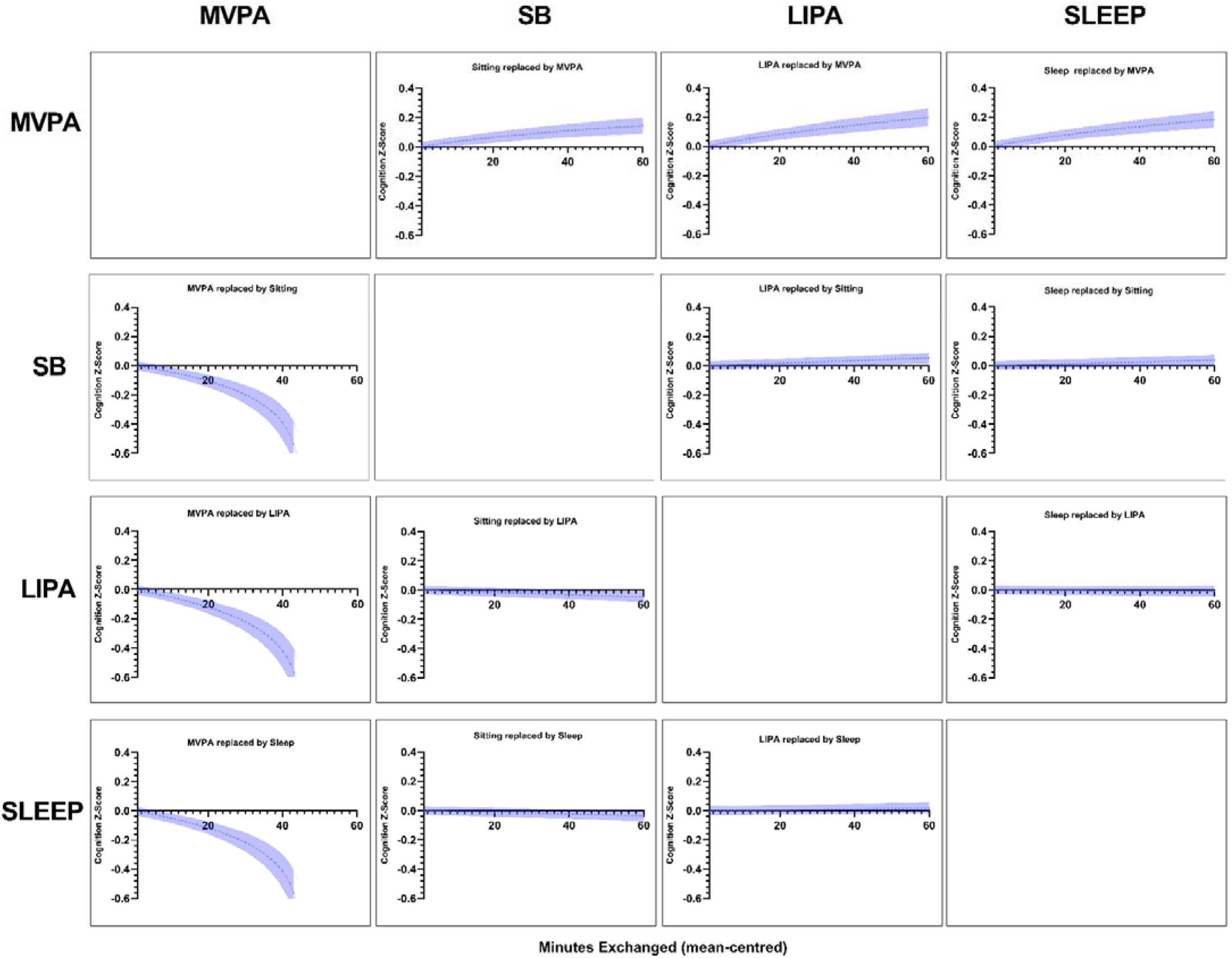
Cognition change following minute-by-minute Isotemporal Substitution between PA types from the mean composition (unadjusted). Relative causal effect on composite cognitive z-scores and 95% confidence intervals of isotemporal substitutions between movement components centred at the mean composition, whilst holding the other two components constant. Substitutions were performed on the first unadjusted ILR model presented in eTable 1 in the supplement.

Replacing LIPA or sleep with SB was also estimated to be favourable for cognition percentile, however, only after 37 minutes of LIPA was replaced by SB (+1.25; 0.03, 2.48; figure 2) or 56 minutes of sleep was replaced by SB (+1.41; 0.01, 2.80).

Most adverse for cognition score however was the reallocation of time away from MVPA. Estimated reductions in cognition percentile were observed when 8 minutes of MVPA was replaced by SB (−1.38; -0.16, -2.59; figure 2), 6 minutes of MVPA was replaced by LIPA (−1.19; -0.01, -2.38) or 7 minutes of MVPA was replaced by sleep (−1.35; -0.15, -2.55).

### Sensitivity Analysis

When analyses were repeated with individual cognitive domains, the relationships between each behaviour relative to all others and memory z-scores proved stronger, and weaker for executive function (eTables 2,3; eFigures 5,6 in the Supplement).

We observed no interaction by sex, but interactions between education and daily movement (*p*<0.005; eTable 4 in the Supplement) and occupational PA and daily movement (*p*<0.01; eTable 5 in the Supplement) were observed. Time reallocation was explored in stratified analysis whereby participants with the highest educational attainment or no formal qualifications appeared to benefit more substantially from reallocating time into MVPA, compared to those in formal education up to age 16 (eFigure 7 in the supplement). Similar trends were seen for those participants in sitting occupations, compared to those with heavy manual jobs (eFigure 8 in the supplement).

## Discussion

Our aim was to examine associations of different components of daily movement with midlife cognition while addressing the co-dependence of movement behaviours. Participant movement profiles were associated with their overall cognition scores. Most notably, MVPA relative to other movement, and SB relative to other behaviours showed positive associations with cognition, with the latter more robust to fuller adjustments. These associations proved stronger for executive function than memory. Our second aim was to explore the relative importance of these associations by modelling time displacements. Permutations of time displacement not involving MVPA (e.g. replacing LIPA with SB) only proved favourable after an unrealistic time reallocation. We observed estimated increases in cognition percentile after 9 minutes of MVPA (above the mean) replaced SB, 7 minutes more MVPA in place of LIPA, or 7 minutes more MVPA in place of sleep. Given it is also the smallest component, the displacement of time away from MVPA quickly proved adverse for cognition percentile. Reallocating 8, 6, and 7 minutes from the mean of MVPA into SB, LIPA or sleep respectively proved sufficient for minor decreases in cognition percentile. Lastly, stratifying by occupation type revealed that participants with more sedentary jobs may show a more substantial cognitive benefit from time being reallocated to MVPA than those participants with more physically demanding employment.

The evidence presented here aligns with previous studies using traditional approaches to pinpoint MVPA as a critical component of daily movement for cognition^35-37,71^. In high income countries, engagement in more vigorous PA is typically reported in higher socioeconomic stratas^72^, where individuals are simultaneously more likely to have a sedentary job^73,74^. However, models proved robust with adjustments for occupational PA, and may suggest a more direct role for MVPA, as opposed to simply a confounder effect. MVPA is typically the most difficult intensity to acquire and was the lowest proportion of the day for all participants in this study. Perhaps partly for this reason, loss of any MVPA time whatsoever appeared detrimental, given loss of each minute of MVPA is many times larger as a proportion of its total than all other daily movements. There are many physiological explanations which may underlie a role for MVPA in supporting cognition including acute increases in cerebral perfusion^75,76^, growth factor release such as brain-derived neurotrophic factor (BDNF)^7^ and even hippocampal neurogenesis^77^. Lastly, MVPA, when attained by structured exercise, involves some degree of self-motivation^78,79^, and often elements of planning, social interaction or teamwork which are all factors considered to be cognitively stimulating^59,80-82^. MVPA relative to other movement proved more strongly associated with executive function, than memory however, suggesting differences in the possible pathways linking PA and different facets of cognition. Future studies exploring the role of MVPA on cognition should attempt to harmonise accelerometer use with an accurate differentiator of physical activity attained from leisure time PA, social and non-social sports, and the associated cognitive demands of each activity.

Despite time reallocation away from SB into MVPA being favourable for cognition, reallocating time from LIPA or sleep into SB was favourably associated with cognition; yet after an unrealistic period. This is contrary to evidence which cite LIPA as favourable and SB as most detrimental to cognition^9,35^. Importantly, our methodology is distinct from these studies which do not wholly address the co-dependent nature of daily movements. One study however, utilising a compositional methodology in a mid-life population also observed this apparent benefit of replacing LIPA and sleep time with SB^37^. Whitaker et al., cites accelerometer inaccuracy specific to waist mounted devices for this relationship. However, we now corroborate this finding using a thigh-mounted device, less susceptible to misclassification of LIPA as SB^57^. Again, we hypothesise that these results may reflect the types of activities one may participate in whilst sedentary. Indeed we may have simply captured typical movement profiles of participants with different lifestyles, rather than any ‘*benefit’* of SB. For instance, participants with more sedentary jobs are likely to have higher educational attainment and participate in cognitively stimulating work^72,74^. Whereas, we posit that participants with manual jobs may spend less time in SB but longer in LIPA. This is corroborated when stratifying by occupational PA, in which we estimated greater improvement in cognition percentile from increasing MVPA in participants in sedentary roles than manual roles. These findings may be explained by distinguishing between SB spent engaging in cognitively stimulating tasks such as reading or working from television-viewing for instance^83,84^. We hypothesise that the optimal balance could therefore lie between spending one’s SB performing cognitively stimulating tasks, whilst also attaining sufficient MVPA in place of any other behaviour including SB itself, given its relative abundance across all participants.

Reallocating time to LIPA was not associated with substantive changes in cognition. LIPA may simply not achieve a threshold intensity to incur any measurable, physiological cognitive benefit, even if providing other cardiometabolic benefits^9^. Greater sleep time was not substantively associated with cognition. Sleep time might have been expected to be highly favourably associated with cognitive performance^40,61,85^. However, sleep in accelerometer studies refers only to time spent in bed and therefore does not perfectly capture biological sleep itself nor its quality. Sleep quality is arguably more strongly associated with cognitive performance than duration^40,85,86^. Differentiating between positive and negative types of SB; sleep duration and sleep quality; and separating leisure time PA from sporting PA, may be avenues of future exploration.

### Strengths and limitations

This study uses objectively measured PA using a robust compositional analysis technique. Use of accelerometers eliminates the risk of PA recall bias and compositional analysis appropriately addresses the co-dependence of each aspect of daily movement. Use of standardised composite cognition scores in this study may provide greater sensitivity than studies which depend on a single measure of global cognition or categorise participants based on stringent test cut points. This is especially true in midlife where the relationship between each measurable aspect of cognition and our daily movements may be more subtle, and given declines in memory and other cognitive abilities through life occur at different rates^87-89^.

Despite these benefits, accelerometer measures do not provide context to each component of movement. SB can involve activities which have positive or negative associations with cognition^83^, for instance constructive leisure activities and working, as well as television viewing. This suggests there may be sub-categories with more nuanced relationships with participant cognition.

Despite use of large sample, the cohort under-represents non-white communities, in which age-related disorders are known to differ in prevalence^90^ limiting the generalisability of these findings to the wider population. Possible sample bias does also exist given participants declining to wear an accelerometer are disproportionately male, smokers and have a health condition compared to those who accepted^57^

Lastly, use of a cross-sectional design limits the inference of the direction of this association. Nonetheless, the risk of reverse causality in these findings may be lessened in this cohort given the younger nature of our cohort. Future such studies must apply these compositional techniques to longitudinal accelerometer-derived movement data encompassing midlife to corroborate these findings.

## Conclusion

The role PA plays in supporting cognition is of increasing interest given the race to address the rise in age-related cognitive decline. This analysis uses compositional methodologies to address the co-dependence of each aspect of daily movement behaviour, and corroborates the role of MVPA in supporting overall cognition, executive function, and memory. The theoretical redistribution of time from MVPA was estimated to be detrimental to all measured facets of cognition after as little as 6 minutes was replaced by other behaviours. SB however, did not appear deleterious relative to other movements, and suggests there may be previously unreported nuance to this relationship.

## Data Availability

Original study protocol and survey documents can be found online at: https://bcs70.info/ and access to this data is available through the UK data service.

## Declarations Acknowledgements

The team would like to pay sincere thanks to the BCS70 participants in this study for their ongoing contribution.

Availability of data and materials: The datasets supporting this article are available in the UK Data Service repository [1970 British Cohort Study: https://beta.ukdataservice.ac.uk/datacatalogue/series/series?id=200001].

## Funding

This study was funded by a British Heart Foundation grant (SP/15/6/31397). JJM is funded by MRC grant (MR/N013867/1). JMB is supported through a British Heart Foundation grant SP/F/20/150002.

## Author Contributions

MH, JB and JJM conceived the study. JJM, JB & SC conducted analysis and interpretation. JJM drafted the manuscript. BJJ, GW and MH examined final analyses and revised several drafts before all authors read and approved the final manuscript.

## Ethics approval and consent to participate

The study has been approved by NRES Committee South East Coast - Brighton & Sussex (Ref 15/LO/1446).). All participants provided ongoing informed consent at age-46 follow-up.

## Availability of data and materials

Original study protocol and survey documents can be found online at: https://bcs70.info/ and access to this data is available through the UK data service: UK Data Service › Series.

## Competing Interests

The authors declare that they have no competing interests or conflicts of interests.

## Additional Materials

Supplementary data for this article is included with this submission.

Additional file 1: Supplementary document of further analysis (.docx file).

## Notes

### Competing Interest Statement

The authors have declared no competing interest.

### Author Declarations

The study has been approved by NRES Committee South East Coast - Brighton & Sussex (Ref 15/LO/1446).). All participants provided ongoing informed consent.

