## Supplement for "Daily movement behaviours and cognition in mid-life: A cross-sectional compositional analysis of the 1970 British Cohort Study"

Supplementary Figure 1. STROBE diagram of sample derivation.

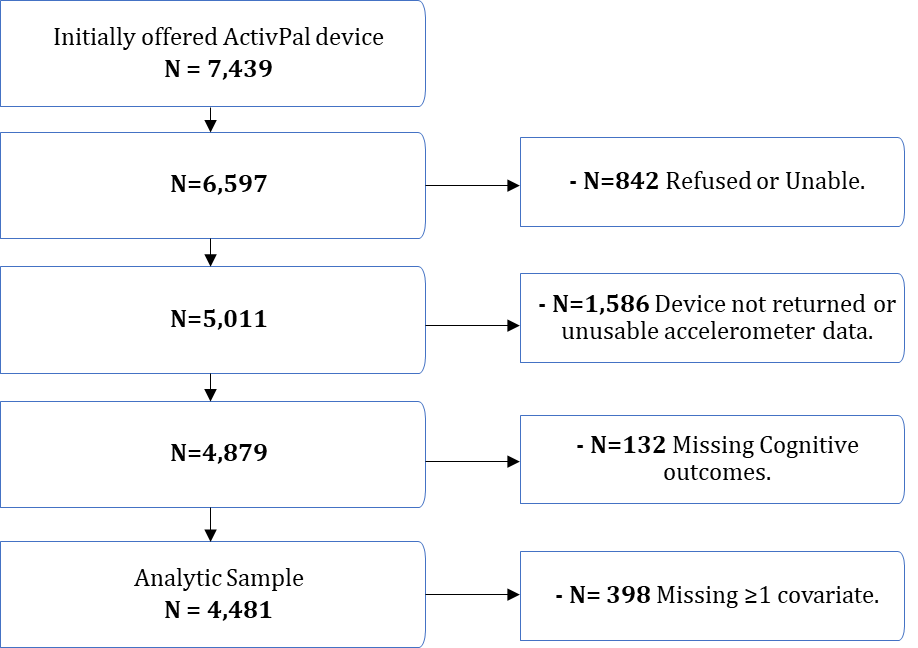

Supplementary Figure 2. Participant average daily movement.

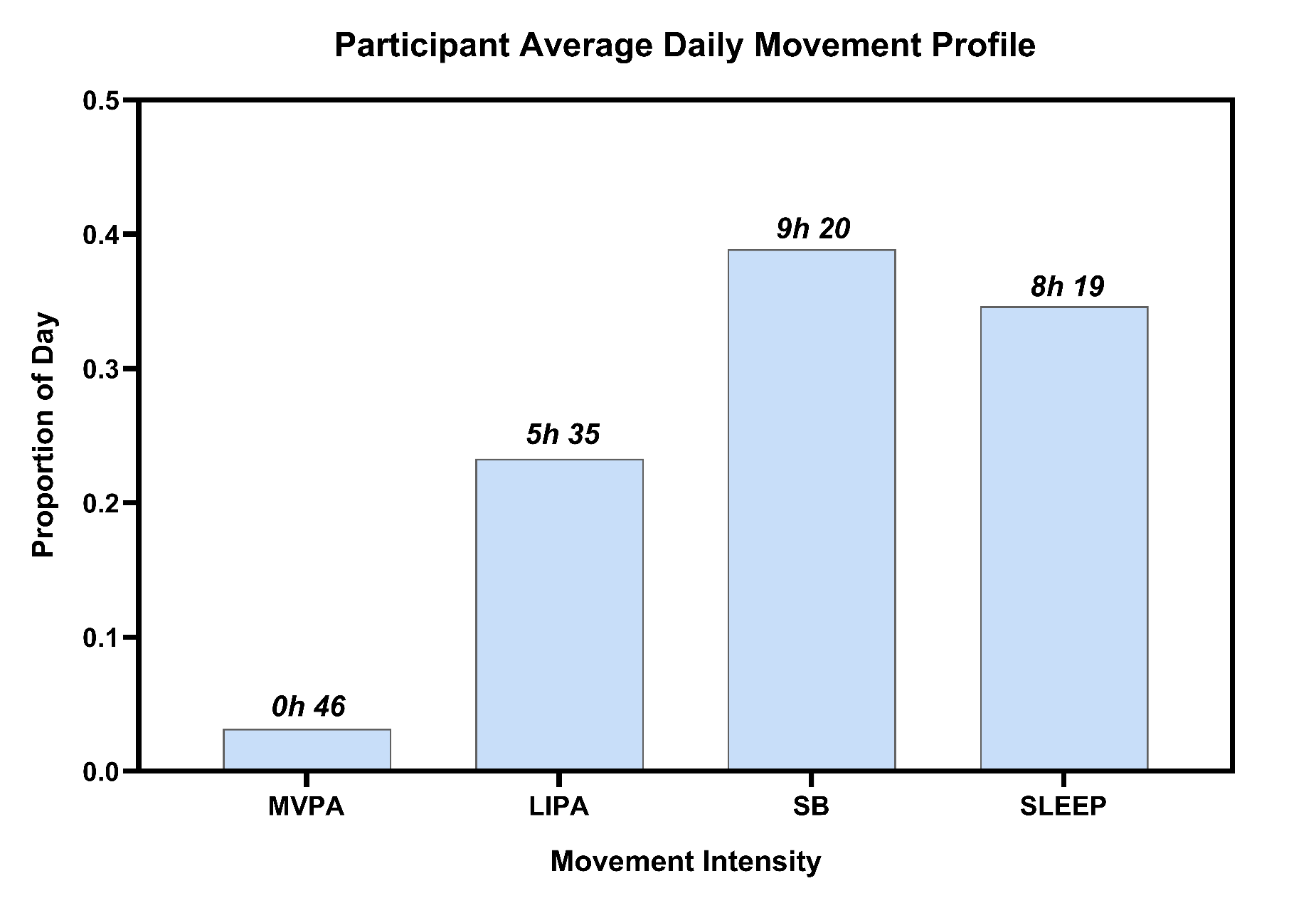

Supplementary Figure 3. Distribution of raw executive function scores (count of letters missed & number of letters process in 2-letter cancellation task; animals named in verbal-fluency task)

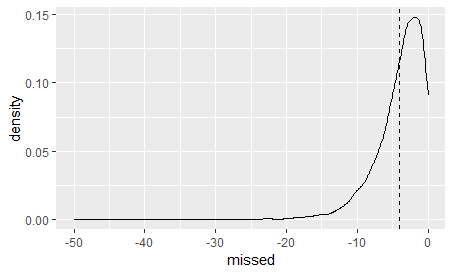

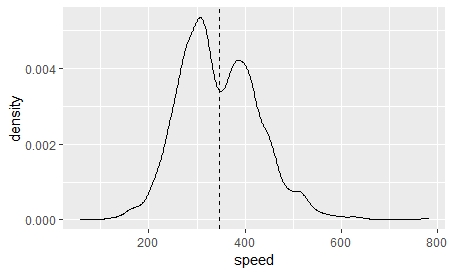

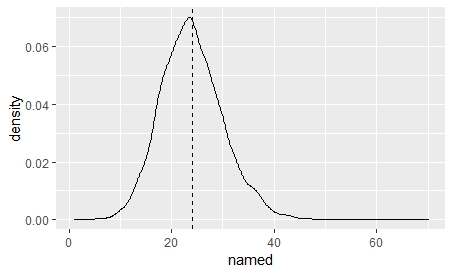

Supplementary Figure 4. Distribution of raw memory scores (*L* – Immediate recall; *R* – Delayed recall)

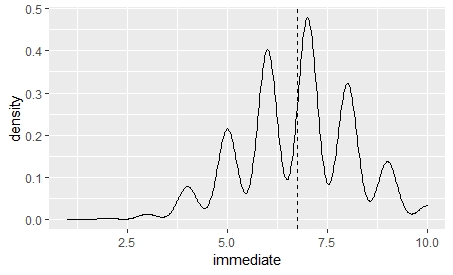

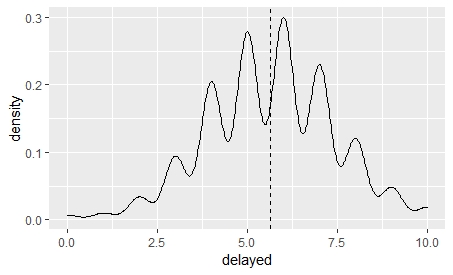

Supplementary Table 1. Estimated coefficients for linear regression model of isometric log ratio (ILR) coordinates of daily time composition and participant composite cognition z-scores

| **Table 3. Estimated coefficients for linear regression model of isometric log ratio (ILR) coordinates of daily time composition and participant composite cognition z-scores.** | | | | | | | | | |
| --- | --- | --- | --- | --- | --- | --- | --- | --- | --- |
| ILR Coordinate | Unadjusted | | | Adjusted for sociodemographic factors including age, sex, education, marital status and socioeconomic status | | | Further adjustment for health and lifestyle factors including disability, BMI, depressive symptoms, smoker status and alcohol consumption. | | |
|  | **Coef.** | **95% CI** | **p-value** | **Coef.** | **95% CI** | **p-value** | **Coef.** | **95% CI** | **p-value** |
| 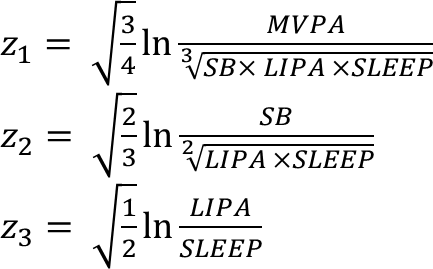 | 0.096 | (0.069 **-** 0.123) | **<0.001** | 0.045 | (0.017 **-** 0.074) | **0.002** | 0.024 | (-0.006 **-** 0.053) | 0.114 |
| 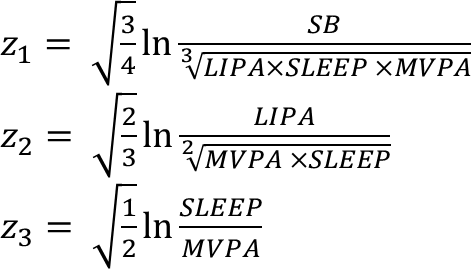 | 0.078 | (0.034 **-** 0.123) | **<0.001** | 0.056 | (0.015 **-** 0.105) | **0.009** | 0.077 | (0.032 **-** 0.122) | **<0.001** |
| 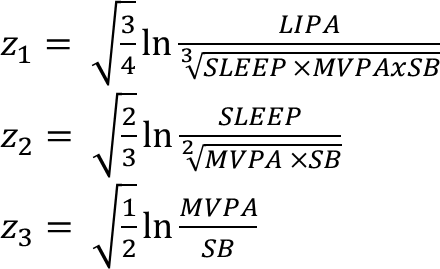 | -0.088 | (-0.177 **-** -0.049) | **<0.001** | -0.025 | (-0.066 **-** 0.016) | 0.230 | -0.016 | (-0.057 **-** 0.025) | 0.438 |
| 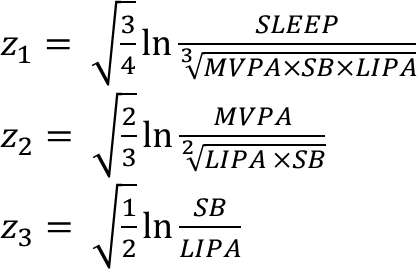 | 0.086 | (-0.142 - -0.030) | **0.003** | -0.080 | (-0.136 **-** -0.024) | **0.005** | -0.084 | (-0.140 **-** -0.029) | **0.003** |

*Coefficients of ILR-transformed coordinates (log-ratio of one behaviour relative to all others) is not immediately interpretable.*

Supplementary Table 2. Analyses repeated for participant memory z-scores only.

| **Table 3. Estimated coefficients for linear regression model of isometric log ratio (ILR) coordinates of daily time composition and participant memory z-scores.** | | | | | | | | | |
| --- | --- | --- | --- | --- | --- | --- | --- | --- | --- |
| ILR Coordinate | Unadjusted | | | Adjusted for sociodemographic factors including, sex, education, marital status and socioeconomic status | | | Further adjustment for health and lifestyle factors including disability, BMI, depressive symptoms, smoker status and alcohol consumption. | | |
|  | **Coef.** | **95% CI** | **p-value** | **Coef.** | **95% CI** | **p-value** | **Coef.** | **95% CI** | **p-value** |
| 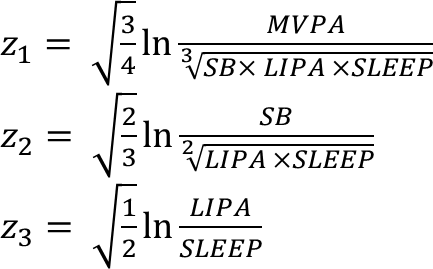 | 0.118 | (0.069 **-** 0.169) | **<0.001** | 0.046 | (-0.003 **-** 0.095) | 0.064 | 0.012 | (-0.038 **-** 0.062) | 0.635 |
| 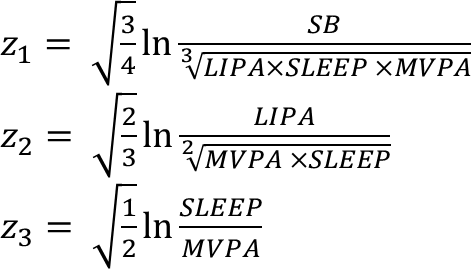 | 0.127 | (0.044 **-** 0.209) | **0.003** | 0.048 | (-0.035 **-** 0.131) | 0.257 | 0.087 | (0.003 **-** 0.171) | **0.042** |
| 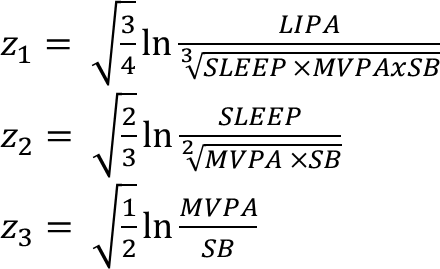 | -0.115 | (-0.189 **-** -0.040) | **0.003** | 0.021 | (-0.055**-** 0.096) | 0.593 | 0.034 | (-0.041 **-** 0.110) | 0.373 |
| 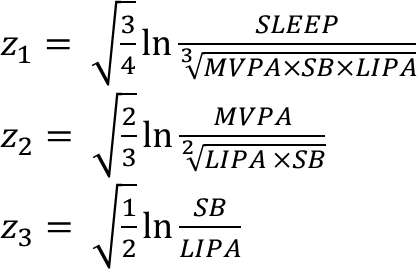 | -0.131 | (-0.236 - -0.027) | **0.014** | -0.115 | (-0.216 **-** -0.014) | **0.026** | -0.117 | (-0.218 **-** 0.016) | **0.023** |

*Coefficients of ILR-transformed coordinates (log-ratio of one behaviour relative to all others) is not immediately interpretable.*

Supplementary Table 3. Analyses repeated for participant executive z-scores only

| **Table 3. Estimated coefficients for linear regression model of isometric log ratio (ILR) coordinates of daily time composition and participant executive function z-scores.** | | | | | | | | | |
| --- | --- | --- | --- | --- | --- | --- | --- | --- | --- |
| ILR Coordinate | Unadjusted | | | Adjusted for sociodemographic factors including age, sex, education, marital status and socioeconomic status | | | Further adjustment for health and lifestyle factors including disability, BMI, depressive symptoms, smoker status and alcohol consumption. | | |
|  | **Coef.** | **95% CI** | **p-value** | **Coef.** | **95% CI** | **p-value** | **Coef.** | **95% CI** | **p-value** |
| 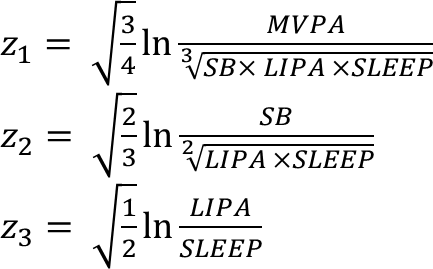 | 0.148 | (0.107 **-** 0.190) | **<0.001** | 0.089 | (0.047 **-** 0.129) | **<0.001** | 0.064 | (0.021 - 0.106) | **0.004** |
| 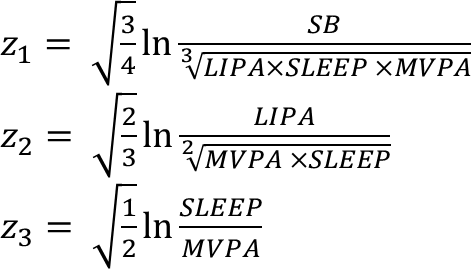 | 0.092 | (0.023 **-** 0.161) | **0.008** | 0.063 | (-0.007 **-** 0.132) | 0.077 | 0.087 | (0.017 **-** 0.158) | **0.015** |
| 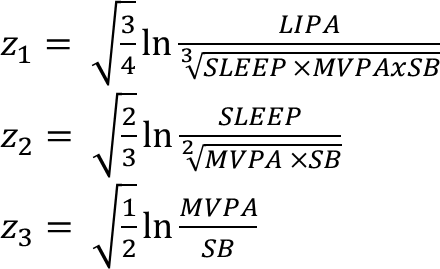 | -0.132 | (-0.194 **-** -0.070) | **<0.001** | -0.054 | (-0.117 **-** 0.009) | 0.095 | -0.051 | (-0.114 **-** 0.012) | 0.113 |
| 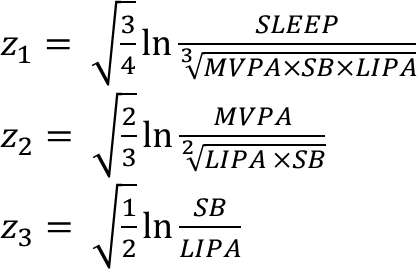 | -0.109 | (-0.196 **-** -0.022) | **0.014** | -0.097 | (-0.182 **-** -0.013) | **0.023** | -0.998 | (-0.184 **-** -0.015) | **0.021** |

*Coefficients of ILR-transformed coordinates (log-ratio of one behaviour relative to all others) is not immediately interpretable.*

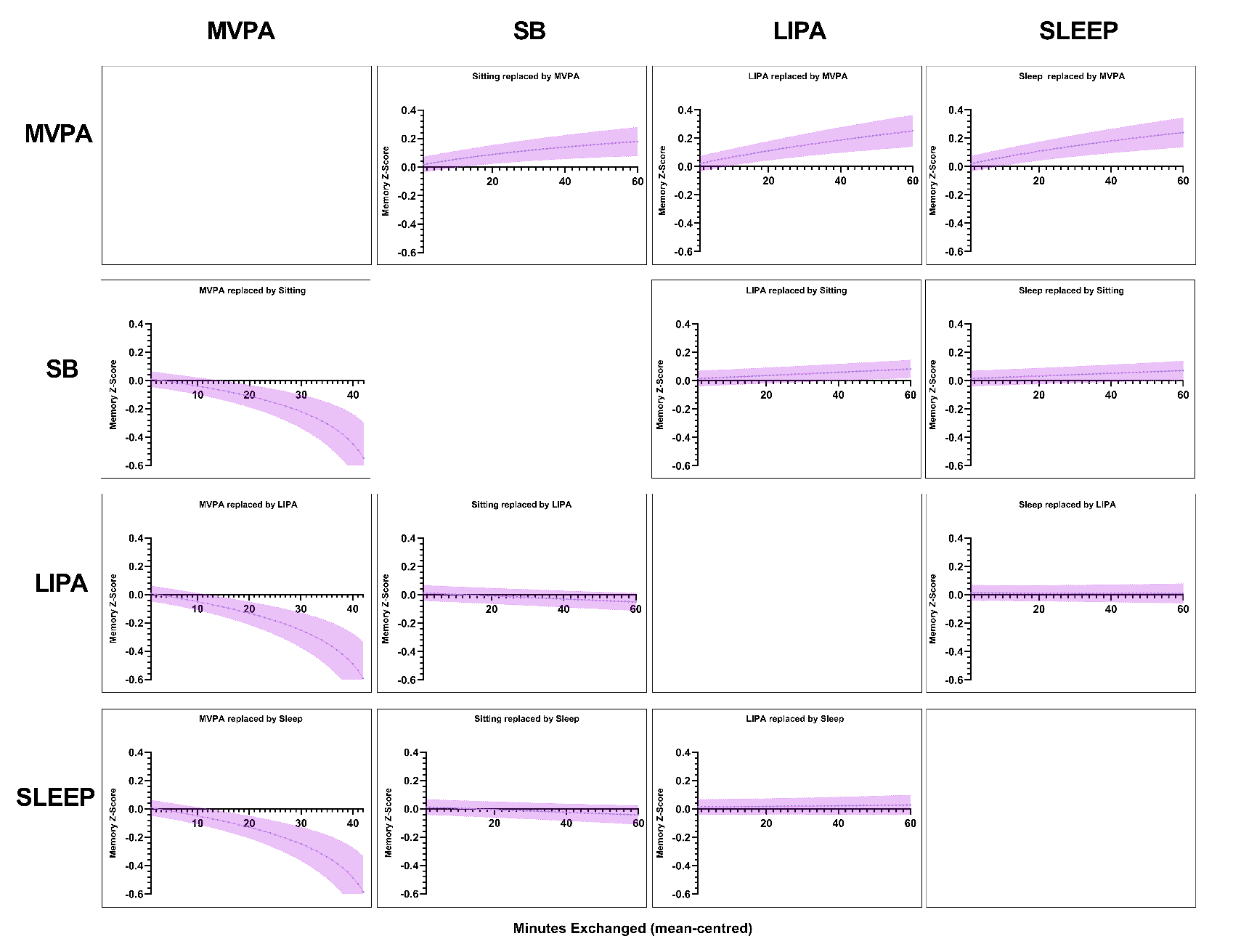
Supplementary Figure 5: Unadjusted isotemporal substitutions between PA intensities and participant memory z-scores.

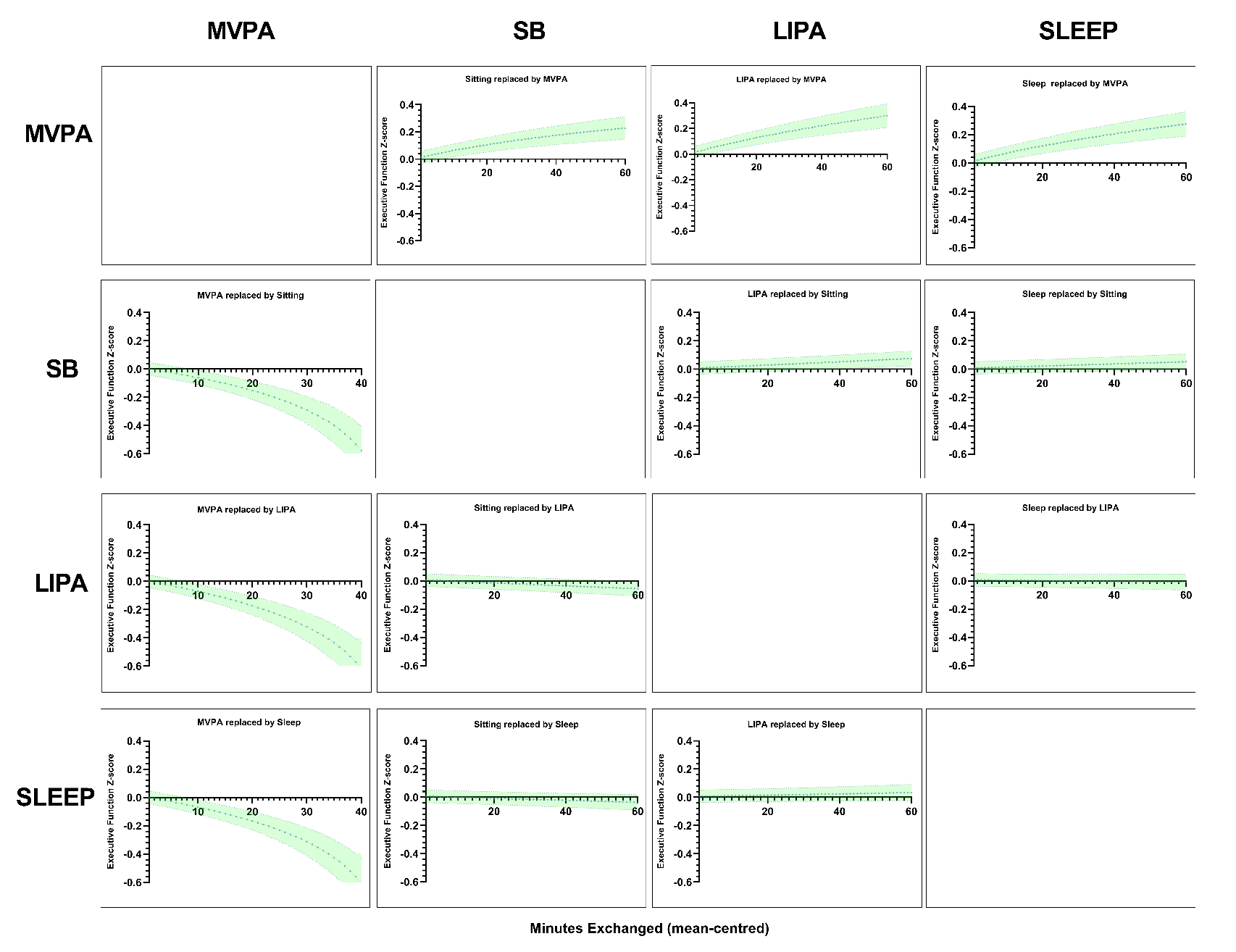
Supplementary Figure 6: Unadjusted isotemporal substitutions between PA intensities and participant executive function.

Supplementary table 4 | Figure 7: Unadjusted education interaction and isotemporal substitutions with composite cognition on outcome stratified by education type (abbreviated).

lm(formula = Cog ~ Zi + Education + Zi:Education)

Residuals:

Min	1Q Median	3Q	Max

-5.7417 -0.6189 0.0282 0.6344 3.1201

Coefficients:] Estimate	Std. Error	t value Pr(>|t|)

(Intercept)	0.014282	0.131665	0.108 0.913629

Zimvpa	0.088717	0.029582	2.999 0.002725 **

Zisedent	0.133827	0.047587	2.812 0.004943 **

ZiLIPA	-0.038694	0.053132 -0.728 0.466504

Education1	-0.373950	0.176911 -2.114 0.034597 *

Education2	-0.107790	0.301752 -0.357 0.720950

Education3	0.646768	0.179253	3.608 0.000312 ***

Education4	0.806157	0.316370	2.548 0.010867 *

Zimvpa:Education1	-0.118925	0.039769 -2.990 0.002803 **

Zisedent:Education1 -0.010957	0.064005 -0.171 0.864080

ZiLIPA:Education1	0.065861	0.071921	0.916 0.359860

Zimvpa:Education2	-0.133008	0.068187 -1.951 0.051169 .

Zisedent:Education2 0.023166	0.109804	0.211 0.832913

ZiLIPA:Education2	-0.022696	0.110759 -0.205 0.837652

Zimvpa:Education3	0.003681	0.040591	0.091 0.927753

Zisedent:Education3 -0.090311	0.068225 -1.324 0.185670

ZiLIPA:Education3	0.090906	0.075885	1.198 0.231012

Zimvpa:Education4	-0.048961	0.072329 -0.677 0.498492

Zisedent:Education4 -0.250937	0.133243 -1.883 0.059734 .

ZiLIPA:Education4	0.026480	0.130436	0.203 0.839139

---

Signif. codes: 0 ‘***’ 0.001 ‘**’ 0.01 ‘*’ 0.05 ‘.’ 0.1 ‘ ’ 1

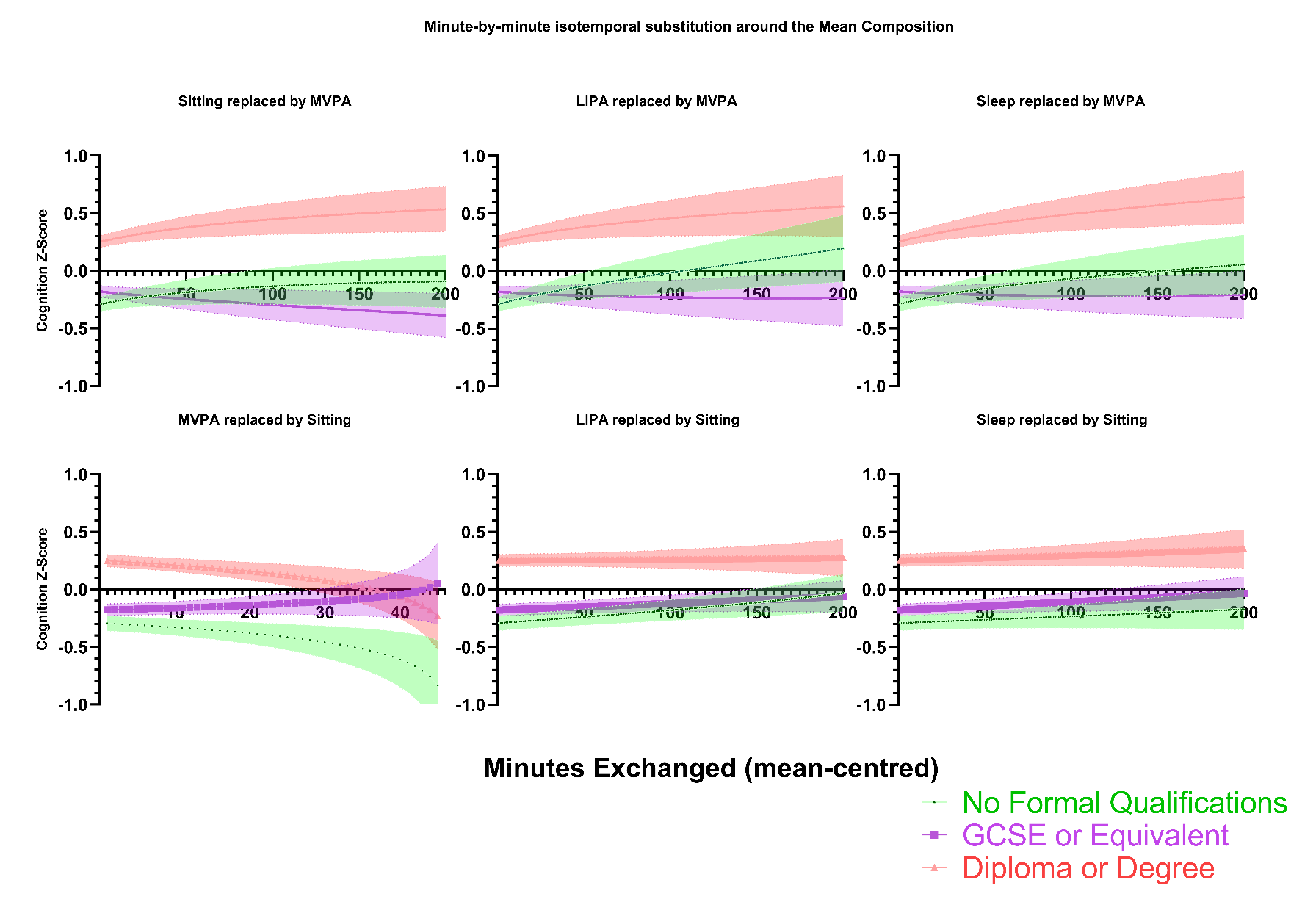

**Figure 7.** Relative causal effect on composite cognitive z-scores and 95% confidence intervals (Stratified by educational level) of isotemporal substitutions between each movement component and SB and MVPA centred at the mean composition. Substitutions were performed on the unadjusted ILR model presented in eTable 1 in the supplement.

Supplementary Table 5 | Figure 8: Unadjusted occupation interaction and isotemporal substitutions with composite cognition on outcome stratified by occupation PA type (abbreviated).

lm(formula = Cog ~ Zi + Occupation_type + Zi:Occupation_type)

Residuals:

Min	1Q Median	3Q	Max

-5.8204 -0.6198 0.0193 0.6577 3.1765

Coefficients: Estimate Std. Error t value Pr(>|t|)

(Intercept)	-0.172104	0.404297 -0.426	0.6704

Zimvpa	0.042916	0.089700	0.478	0.6324

Zisedent	-0.123092	0.164447 -0.749	0.4542

ZiLIPA	-0.066044	0.167380 -0.395	0.6932

Occupation_type1	0.689221	0.415985	1.657	0.0976 .

Occupation_type2	0.329965	0.447994	0.737	0.4614

Occupation_type3	0.051099	0.426484	0.120	0.9046

Occupation_type4	-0.422516	0.510941 -0.827	0.4083

Zimvpa:Occupation_type1	0.028922	0.092180	0.314	0.7537

Zisedent:Occupation_type1 0.154173	0.169352	0.910	0.3627

ZiLIPA:Occupation_type1	0.137075	0.172384	0.795	0.4266

Zimvpa:Occupation_type2	-0.008646	0.100697 -0.086	0.9316

Zisedent:Occupation_type2 0.214331	0.177580	1.207	0.2275

ZiLIPA:Occupation_type2	0.063925	0.182997	0.349	0.7269

Zimvpa:Occupation_type3	-0.011420	0.095070 -0.120	0.9044

Zisedent:Occupation_type3 0.190386	0.172439	1.104	0.2696

ZiLIPA:Occupation_type3	0.054388	0.177441	0.307	0.7592

Zimvpa:Occupation_type4	-0.071969	0.113175 -0.636	0.5249

Zisedent:Occupation_type4 0.321077	0.191153	1.680	0.0931 .

ZiLIPA:Occupation_type4	0.167670	0.214274	0.783	0.4340

---

Signif. codes: 0 ‘***’ 0.001 ‘**’ 0.01 ‘*’ 0.05 ‘.’ 0.1 ‘ ’ 1

Residual standard error: 0.9749 on 3807 degrees of freedom

Multiple R-squared: 0.04887,	Adjusted R-squared: 0.04412

F-statistic: 10.29 on 19 and 3807 DF, p-value: < 2.2e-16

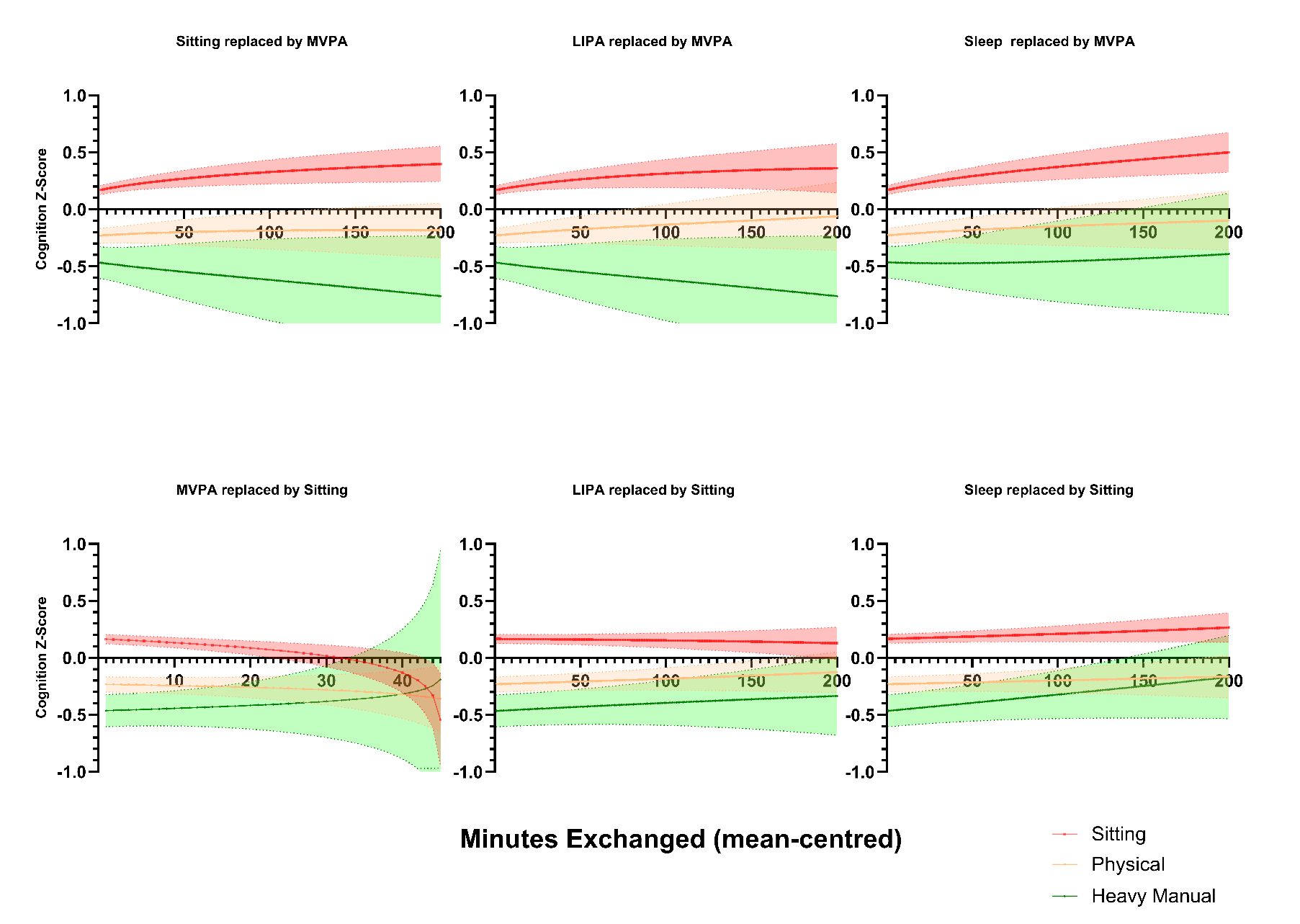

**Figure 8.** Relative causal effect on composite cognitive z-scores and 95% confidence intervals (Stratified by occupational PA level) of isotemporal substitutions between each movement component and SB and MVPA centred at the mean composition. Substitutions were performed on the unadjusted ILR model presented in eTable 1 in the supplement.
